# Preoperative COVID-19 Vaccination is Associated with Decreased Perioperative Mortality after Major Vascular Surgery

**DOI:** 10.1101/2024.03.11.24304133

**Authors:** Molly Ratner, Karan Garg, Heepeel Chang, Anjali Nigalaye, Steven Medvedosky, Glenn Jacobowitz, Jeffrey J Siracuse, Virendra Patel, Marc Schermerhorn, Charles DiMaggio, Caron B Rockman

**Affiliations:** Division of Vascular and Endovascular Surgery, Department of Surgery, New York University Langone Medical Center, New York, NY; Division of Vascular Surgery, Department of Surgery, Westchester Medical Center, Valhalla, NY; Division of Hospitalist Medicine, Department of Medicine, Mount Sinai Beth Israel Hospital, New York, NY; Division of Vascular and Endovascular Surgery, Department of Surgery, Boston Medical Center, Boston, Massachusetts; Division of Vascular and Endovascular Interventions, Department of Surgery, New York Presbyterian-Columbia University Irving Medical Center, New York, NY; Division of Vascular and Endovascular Surgery, Department of Surgery, Beth Israel Deaconess Medical Center, Boston, Massachusetts

**Author notes:** **Pre-publication Corresponding Author:** Molly Ratner, MD, Division of Vascular and Endovascular Surgery, Department of Surgery, New York University Langone Medical Center, NYU Grossman School of Medicine, 550 First Avenue, New York, NY 10016, USA. Drs Molly Ratner and Karan Garg contributed equally to this manuscript.

## Abstract

**Background:** The objective of this study was to examine the effect of COVID-19 vaccination on perioperative outcomes after major vascular surgery.

**Methods:** This is a multicenter retrospective study of patients who underwent major vascular surgery between December 2021 through August 2023. The primary outcome was all-cause mortality within 30 days of index operation or prior to hospital discharge. Multivariable models were used to examine the association between vaccination status and the primary outcome.

**Results:** Of the total 85,424 patients included, 19161 (22.4%) were unvaccinated. Unvaccinated patients were younger compared to vaccinated patients (mean age 68.44 +/− 10.37 years vs 72.11 +/− 9.20 years, p <.001) and less likely to have comorbid conditions, including hypertension (87.2% vs 89.7%, p <.001), congestive heart failure (14.5% vs 15.9%, p <.001), chronic obstructive pulmonary disease (35.7% vs 36.3, p <.001) and renal failure requiring hemodialysis (1.4% vs 1.7%, p = .005). After risk factor adjustment, vaccination was associated with decreased mortality (OR 0.7, 95% CI 0.62 - 0.81, p <.0001). Stratification by procedure type demonstrated that vaccinated patients had decreased odds of mortality after open AAA (OR 0.6, 95% CI 0.42-0.97, p = 0.03), EVAR (OR 0.6, 95% CI 0.43-0.83, p 0.002), CAS (OR 0.7, 95% CI 0.51-0.88, p = 0.004) and infra-inguinal lower extremity interventions (OR 0.7, 95% CI 0.48-0.96, p = 0.03).

**Conclusions:** COVID-19 vaccination is associated with reduced perioperative mortality in patients undergoing vascular surgery. This association is most pronounced for patients undergoing aortic aneurysm repair, carotid stenting and infrainguinal bypass.

## Introduction

Although primarily an infection of the respiratory tract, Coronavirus Disease 2019 (COVID-19) generates a systemic inflammatory state leading to dysregulation of and injury to multiple organ systems,^1–5^ including the vasculature.^6,7^ In the perioperative period, it has been proposed that the exaggerated immune response elicited by both surgical trauma and severe acute respiratory syndrome coronavirus 2 (SARS-CoV-2) may be responsible for the increased risk of perioperative mortality and morbidity in patients with active or recent infections.^8,9^ These findings have prompted groups like the American Society of Anesthesiologists to recommend delaying elective surgery in patients with COVID-19.^10^ However, as the pandemic enters a new phase with widespread population immunity in the setting of vaccine development and/or prior infection, risk stratification will need to reflect these changes.

COVID-19 vaccination has been associated with improved outcomes, including decreased mortality, in patients undergoing various surgical procedures, such as cardiovascular, gastrointestinal operations and orthopedic surgeries.^11–13^ However, there is a paucity of literature evaluating the effect of COVID-19 vaccination on perioperative outcomes after major vascular surgery, specifically. In this retrospective cohort study, we utilized the Society for Vascular Surgery Vascular Quality Initiative (SVS-VQI) database to examine national trends in perioperative mortality after major vascular surgery in the post-vaccination stage of the pandemic.

## Methods

### Data Source

This study utilized the SVS-VQI, a prospectively maintained set of 14 registries containing standardized data collected from over 1000 institutions across the United States, Canada and Singapore. The VQI stores demographic, perioperative and postoperative data on major vascular procedures for the purpose of improving surgical outcomes. Approval was obtained from the SVS VQI Patient Safety Organization prior to analysis. The study was exempt from Institutional Board Review approval and individual patient consent was waived given its retrospective nature.

### Study Population and Definitions

The SVS-VQI was queried from the start of COVID vaccination data collection on December 16^th^, 2021, through its termination on August 31, 2023. Patients were included if they underwent a “major” vascular surgery procedure (i.e. those performed mainly in a non-ambulatory setting), including aortic aneurysm repair (open vs endovascular abdominal/thoracic), carotid artery endarterectomy or stenting and open peripheral revascularization. Endovascular peripheral vascular interventions (PVI) were excluded given the shift towards performing these procedures in an outpatient or ambulatory care setting.^14^

The primary variable of interest was COVID vaccination status. Vaccinated patients included those who had completed the full vaccine series by the time of surgery or had received at least one vaccine with the intention of completing the series. Baseline characteristics, such as demographic and medical history, were collected and included: age, sex, race, smoking status, hypertension, hyperlipidemia, diabetes, coronary artery disease (CAD), congestive heart failure (CHF), chronic obstructive pulmonary disease (COPD), cerebrovascular disease (CVD), renal failure requiring hemodialysis (HD) and preoperative medication utilization. The urgency of the case (i.e. elective vs urgent/emergent) was additionally considered a potential confounder.

### Outcomes

The primary outcome was perioperative all-cause mortality, defined as occurring either prior to discharge from the hospital or within 30 days of index operation. The secondary endpoints were surgery-specific perioperative mortality.

### Statistical Analysis

All analyses were conducted in the R statistical software program. Comparisons of continuous variables were based on t-tests and categorical variables were compared using Chi Square tests as part of the CreatTableOne function of the R tableone package. Unadjusted odds ratios (ORs) for the association of COVID vaccination with perioperative mortality were calculated using the epitab function of the R epitools package. Separate multivariable logistic regression models for each type of procedure (open repair, EVAR, TEVAR, carotid endarterectomy, carotid artery stenting, infrainguinal and suprainguinal revascularization) comparing the association of COVID vaccination with perioperative mortality were adjusted for confounding by age, gender, hypertension, diabetes, race, ethnicity, pre-operative history of smoking and whether the procedure was elective or urgent. A single model with an additional control variable for the type of procedure was run for the full combined data. An additional multilevel model which set the intercept to vary by procedure type was created to establish a baseline risk for mortality risk for each procedure cohort. Results of the multivariable and multilevel models are presented as ORs with associated p-values and 95% confidence intervals (CIs).

## Results

### Baseline Characteristics

Of the total 85,424 patients identified, 19161 (22.4%) were unvaccinated at the time of index operation. Unvaccinated patients were significantly younger compared to vaccinated patients (mean age 68.44 +/− 10.37 years vs 72.11 +/− 9.20 years, p <0.001). The majority of patients in both cohorts were male (65.0% vs 65.6%, p = 0.34) and white, although black patients were over-represented in the unvaccinated group (8.3% vs 7.6%, p = 0.001). Unvaccinated patients were more likely to be current smokers (39.4% vs 25.5%, p <0.001) compared to vaccinated patients. Unvaccinated patients were less likely to have a history of hypertension (87.2% vs 89.7%, p <0.001), CHF (14.5% vs 15.9%, p <0.001), COPD (35.7% vs 36.3, p <0.001) and HD dependence (1.4% vs 1.7%, p = 0.005). Unvaccinated patients were additionally less likely to be taking medications for cardiovascular risk reduction, including aspirin (75.2% vs 77.9%, p <0.001), statins (77.9% vs 84.7%, p <0.001) and beta-blockers (44.9% vs 50.4%, p <0.001). Unvaccinated patients were more likely to have a positive pre-or postoperative positive COVID test, although the overall incidence was low (1.9% vs 1.1%, p <0.001). Unvaccinated patients also less frequently underwent elective procedures (76.2% vs 83.5%, p <0.001). See Table I for a full list of baseline characteristics.

**Table I:** Baseline comorbidities and characteristics of unvaccinated and vaccinated patients undergoing major vascular surgery.

|  | Unvaccinated (n = 19161) | Vaccinated (n = 66263) | P Value |
| --- | --- | --- | --- |
| Age (mean +/- SD) | **68.44 +/- 10.37** | **72.11 +/- 9.20** | **<0.001** |
| Male | 12461 (65.0%) | 43479 (65.6%) | 0.14 |
| Race |  |  |  |
| White | 16191 (84.5%) | 56359 (85.1%) | 0.06 |
| Black | **1584 (8.3%)** | **5009 (7.6%)** | **.001** |
| Current Smoker | **7245 (37.8%)** | **16896 (25.5%)** | **<0.001** |
| Former Smoker | **7260 (37.9%)** | **33248 (50.2%)** | **<.001** |
| Hypertension | **16700 (87.2%)** | **59442 (89.7%)** | **<0.001** |
| Diabetes | 6957 (36.3%) | 24225 (36.5%) | 0.53 |
| CAD | 6422 (33.5%) | 22601 (34.1%) | 0.13 |
| CABG | **2944 (15.4%)** | **11438 (17.3%)** | **<0.001** |
| PCI | **4217 (22.0%)** | **15381 (23.2%)** | **0.001** |
| CHF | **2771 (14.5%)** | **10523 (15.9%)** | **<0.001** |
| COPD | **5423 (35.7%)** | **17633 (36.3%)** | **<0.001** |
| Hemodialysis | **270 (1.4%)** | **1129 (1.7%)** | **0.005** |
| Preoperative Medications |  |  |  |
| Aspirin | **14413 (75.2%)** | **51610 (77.9%)** | **<0.001** |
| Statin | **14925 (77.9%)** | **56021 (84.7%)** | **<0.001** |
| ACE inhibitor | **9546 (49.8%)** | **35075 (53.0%)** | **<0.001** |
| P2Y<sub>12</sub>-Inhibitor | **9470 (49.4%)** | **32123 (48.5%)** | **0.02** |
| Anticoagulation | **2511 (13.1%)** | **9698 (14.6%)** | **<0.001** |
| Beta Blocker | **8606 (44.9%)** | **33354 (50.4%)** | **<0.001** |
| Elective Status | **14593 (76.2%)** | **55252 (83.5%)** | **<0.001** |
| Positive COVID Status | **358 (1.9%)** | **734 (1.1%)** | **<0.001** |
| Perioperative Mortality | **360 (1.8%)** | **761 (1.1%)** | **<0.001** |

CAD, coronary artery disease; CABG, coronary artery bypass graft; PCI, percutaneous coronary intervention; CHF, congestive heart failure; COPD, chronic obstructive pulmonary disease; ACE-inhibitor, angiotensin converting enzyme inhibitor
Bold faced p values indicate significance.

### Multivariable Model

After adjusting for age, gender, race, ethnicity, smoking status, hypertension, diabetes, coronary artery disease and dialysis, vaccinated status was associated with decreased mortality (OR 0.7, 95% CI 0.62 - 0.81, p <0.001). A multilevel model with the same variables in the multivariable logistic model with a random intercept by procedure type returned the same estimate for the effect of the vaccine (OR 0.7, 95% CI 0.62 −0.81). After stratifying by procedure type, vaccinated patients had decreased odds of mortality after open AAA (OR 0.6, 95% CI 0.42-0.97, p = 0.03), EVAR (OR 0.6, 95% CI 0.43-0.83, p= 0.002), CAS (OR 0.7, 95% CI 0.51-0.88, p = 0.004) and infra-inguinal lower extremity interventions (OR 0.7, 95% CI 0.48-0.96, p = 0.03). However, vaccinated status did not significantly reduce the odds of perioperative death in patients who underwent TEVAR, CEA or supra-inguinal lower extremity intervention. See Figure 1 for the Forest Plot.

**Figure 1:**
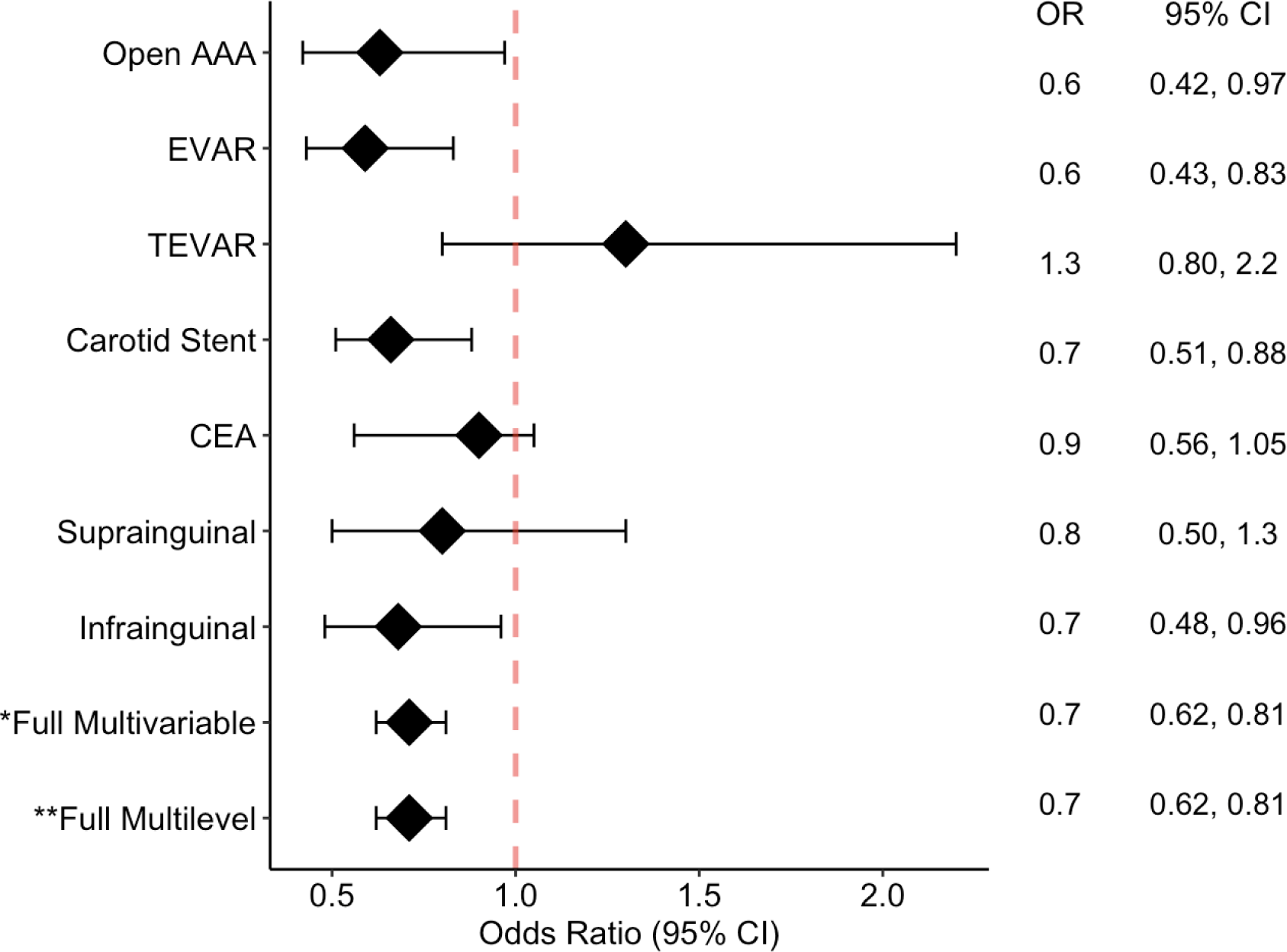
Forest Plot Evaluating COVID Vaccination Status and Mortality

### Procedure Type

#### AORTIC SURGERY (TABLE II)

**Table II:**
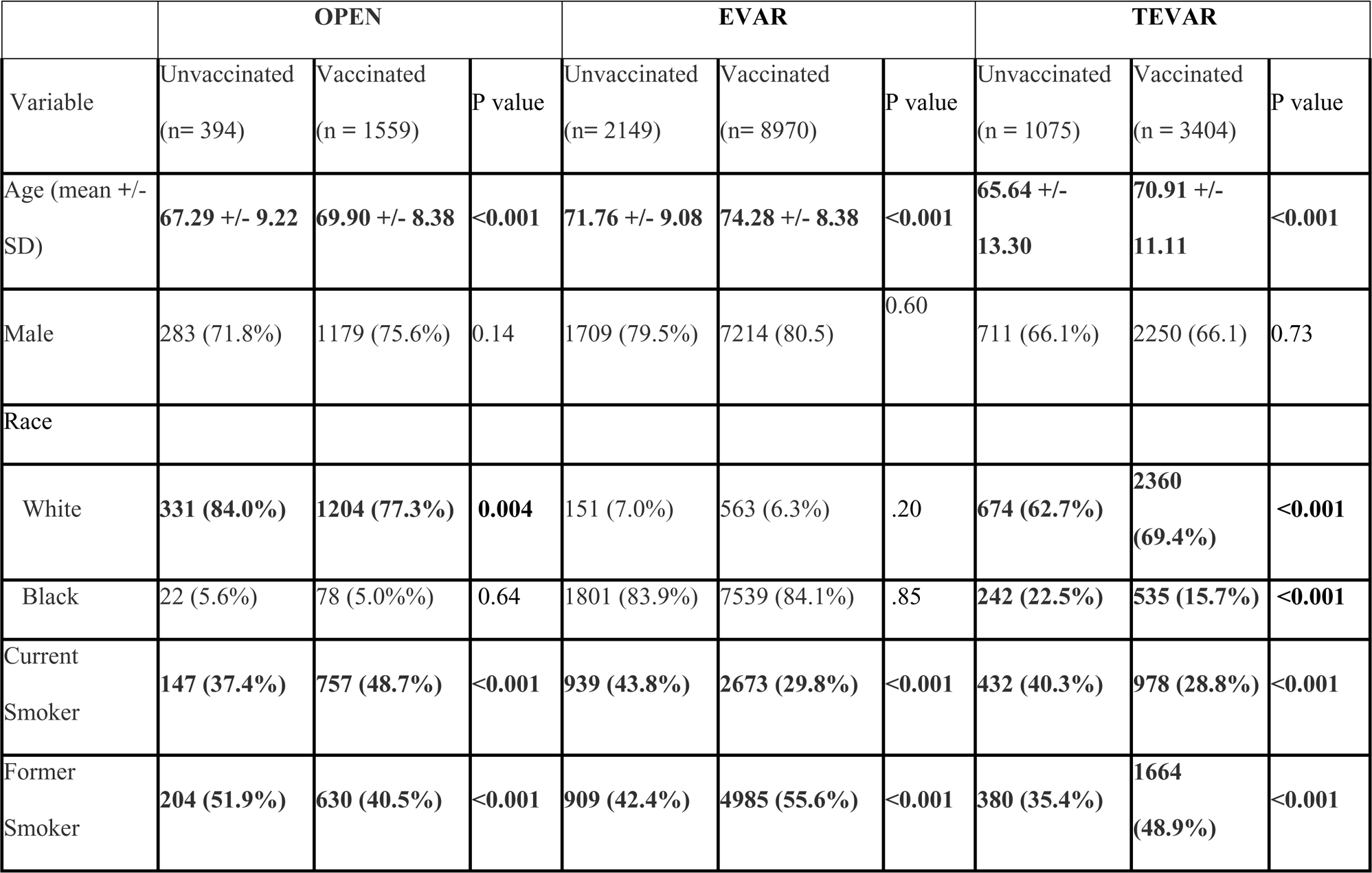
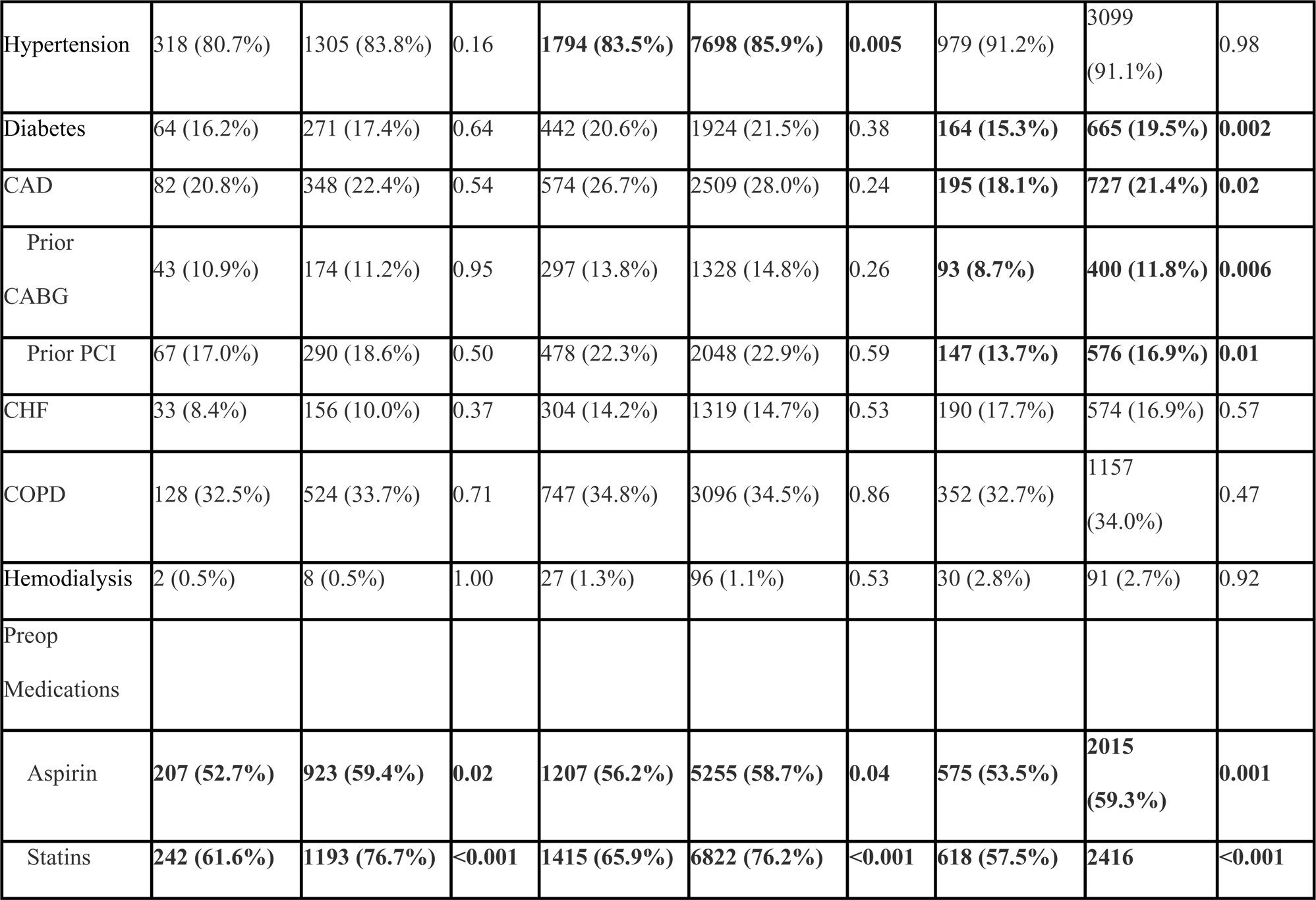
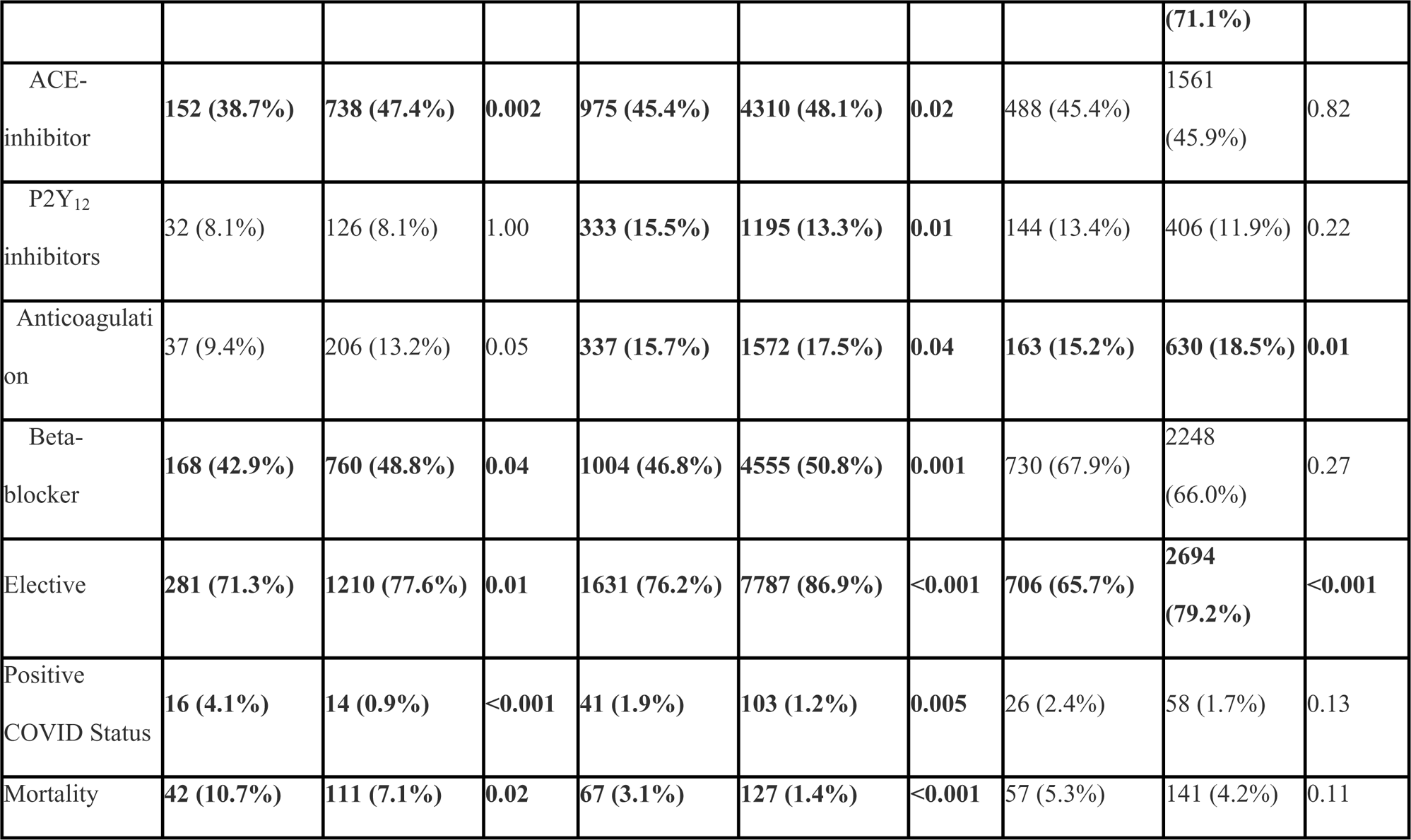
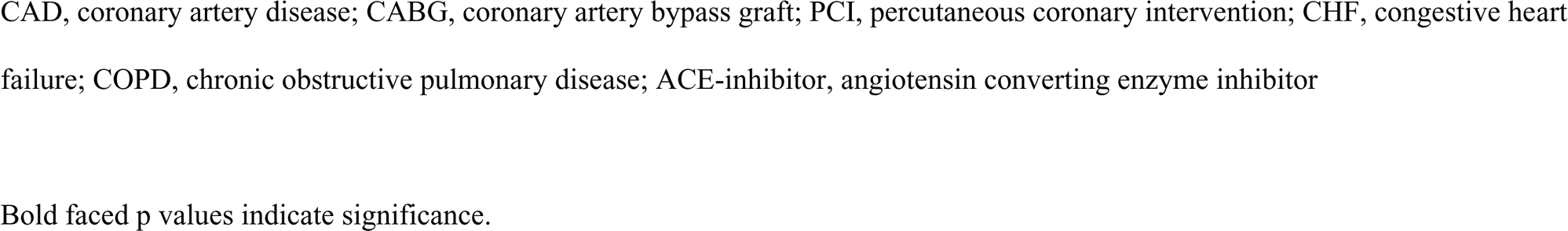
Baseline comorbidities and characteristics of unvaccinated and vaccinated patients undergoing aortic surgery.

Similar to the overall cohort population, unvaccinated patients undergoing OAR, EVAR and TEVAR were significantly younger than their vaccinated counterparts. Unlike the general cohort, baseline comorbidities were similar between vaccinated and unvaccinated patients with a few notable exceptions. Vaccinated patients who underwent TEVAR were more likely to have preoperative diabetes (19.5% vs 15.3%, p = 0.002) and CAD (21.4% vs 18.1%, p = 0.02) compared to the unvaccinated cohort. For all patients undergoing aortic surgery, vaccinated patients were more likely to be on aspirin and statins. In this cohort, unvaccinated patients were significantly less likely than vaccinated patients to undergo surgery electively. Vaccinated patients who underwent OAR and EVAR were less likely to have a positive covid diagnosis (0.9% vs 4.1%, p = 0.02; 1.4% vs 3.1%, p <0.001) and perioperative mortality was significantly lower compared to unvaccinated patients (7.1% vs 10.7%, p = 0.02; 3.1% vs 1.4%, p <0.001). There was no significant difference in covid status or perioperative mortality between unvaccinated and vaccinated patients who underwent TEVAR.

### Carotid Artery Revascularization (Table III)

**Table III:**
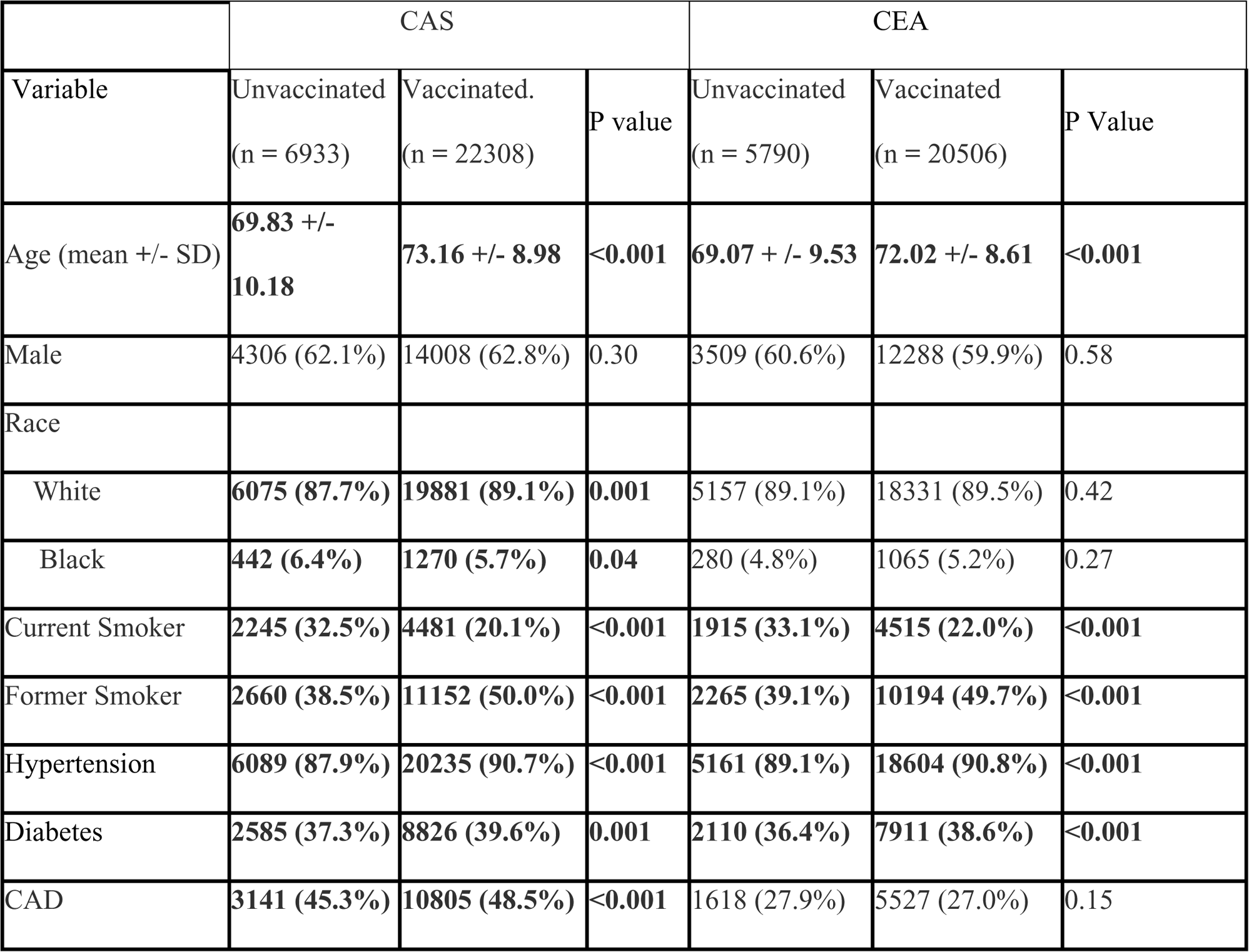
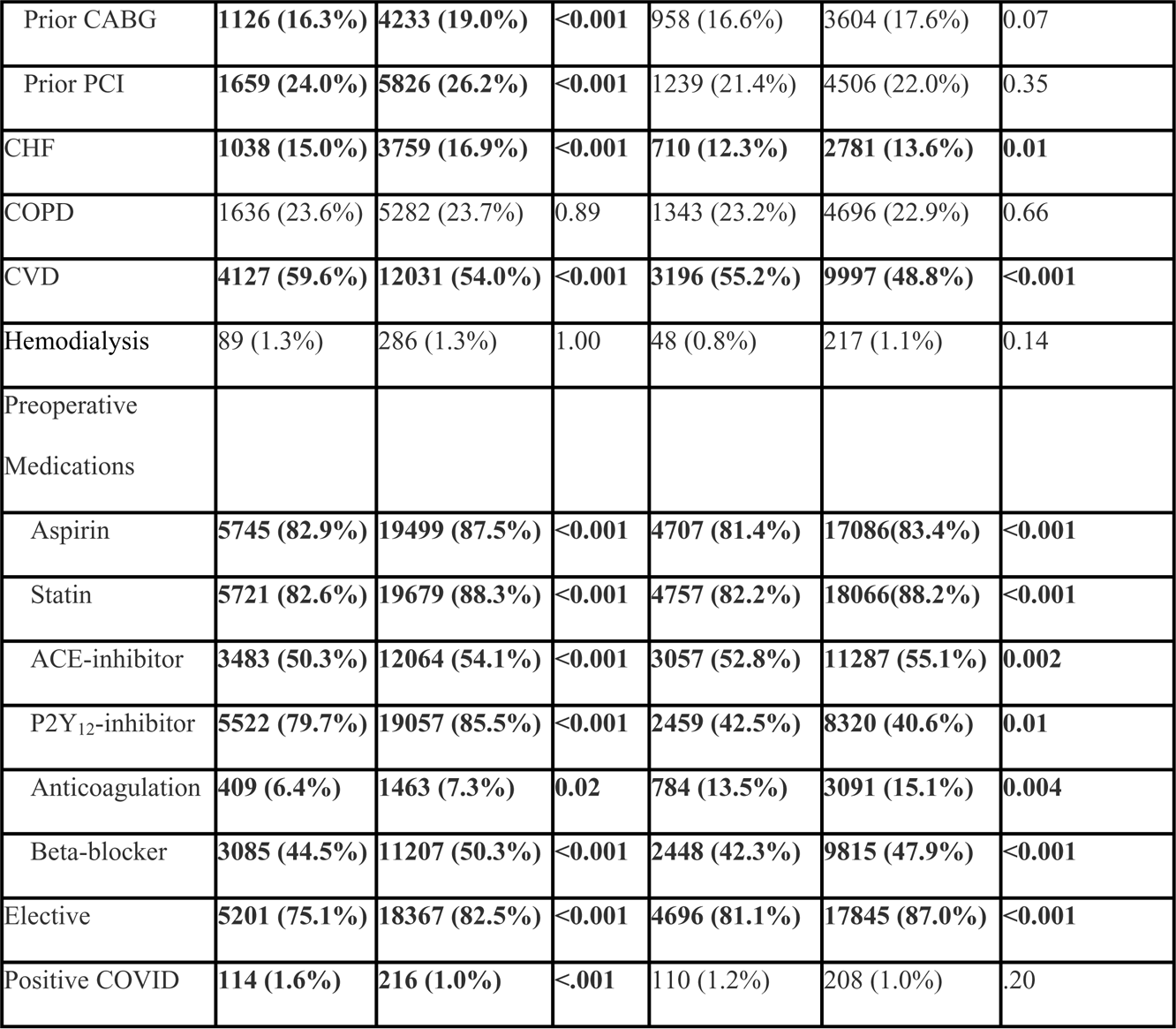
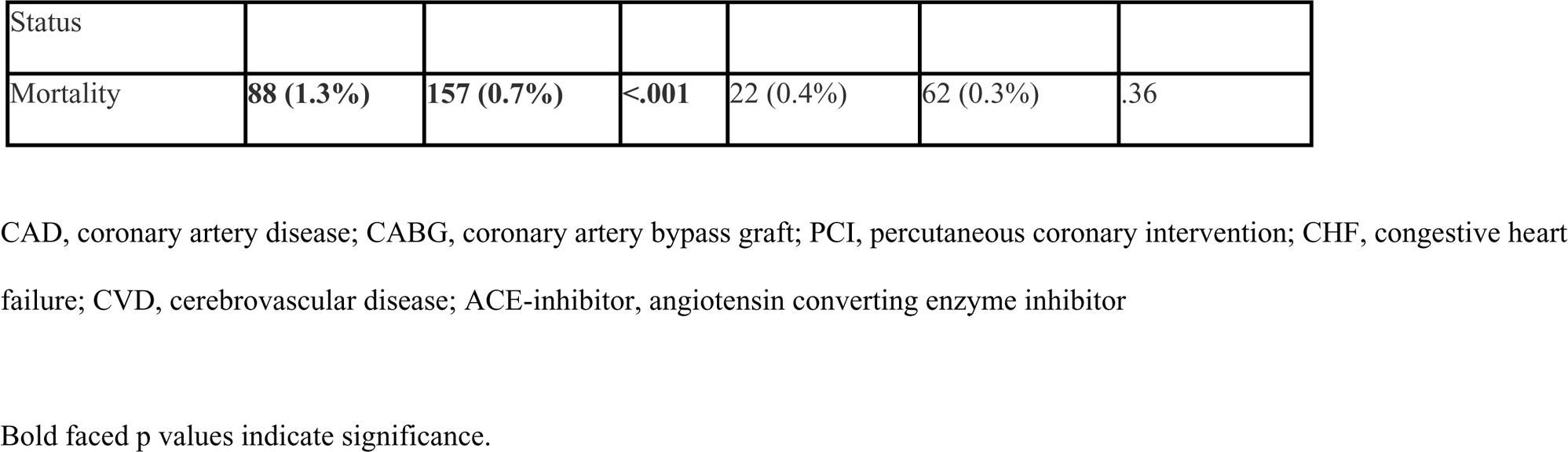
Baseline comorbidities and characteristics of unvaccinated and vaccinated patients undergoing carotid artery revascularization.

Similar to the overall patient cohort, vaccinated patients undergoing CAS and CEA were more likely to have a variety of medical comorbidities including hypertension, diabetes, and CHF. Notably, however, they were less likely than unvaccinated patients to have a history of CVD (54% vs 59.6%, p <0.001; 48.8% vs 55.2%, p <0.001). Vaccinated patients who underwent carotid revascularization were more likely to be on an aspirin, statin, ACE-inhibitor, or beta-blocker. Vaccinated patients who underwent CAS were less likely to have a positive covid diagnosis (1.0% vs 1.6%, p <.001) and peri-operative mortality was significantly reduced compared to the unvaccinated cohort (0.7% vs 1.3%, p <0.001). There was no significant difference in covid status or perioperative mortality between unvaccinated and vaccinated patients who underwent CEA.

### Peripheral Revascularization (Table IV)

**Table IV:**
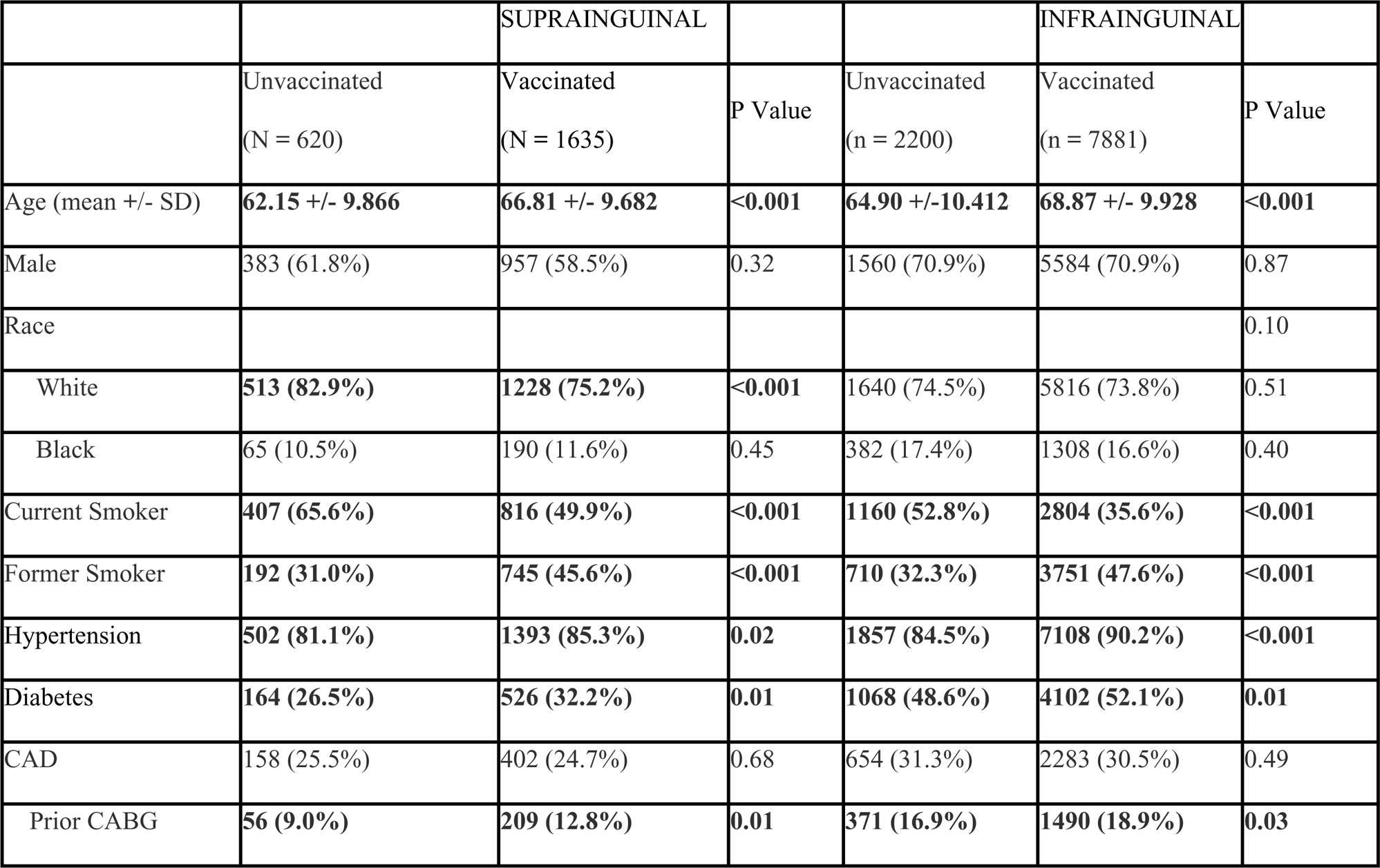
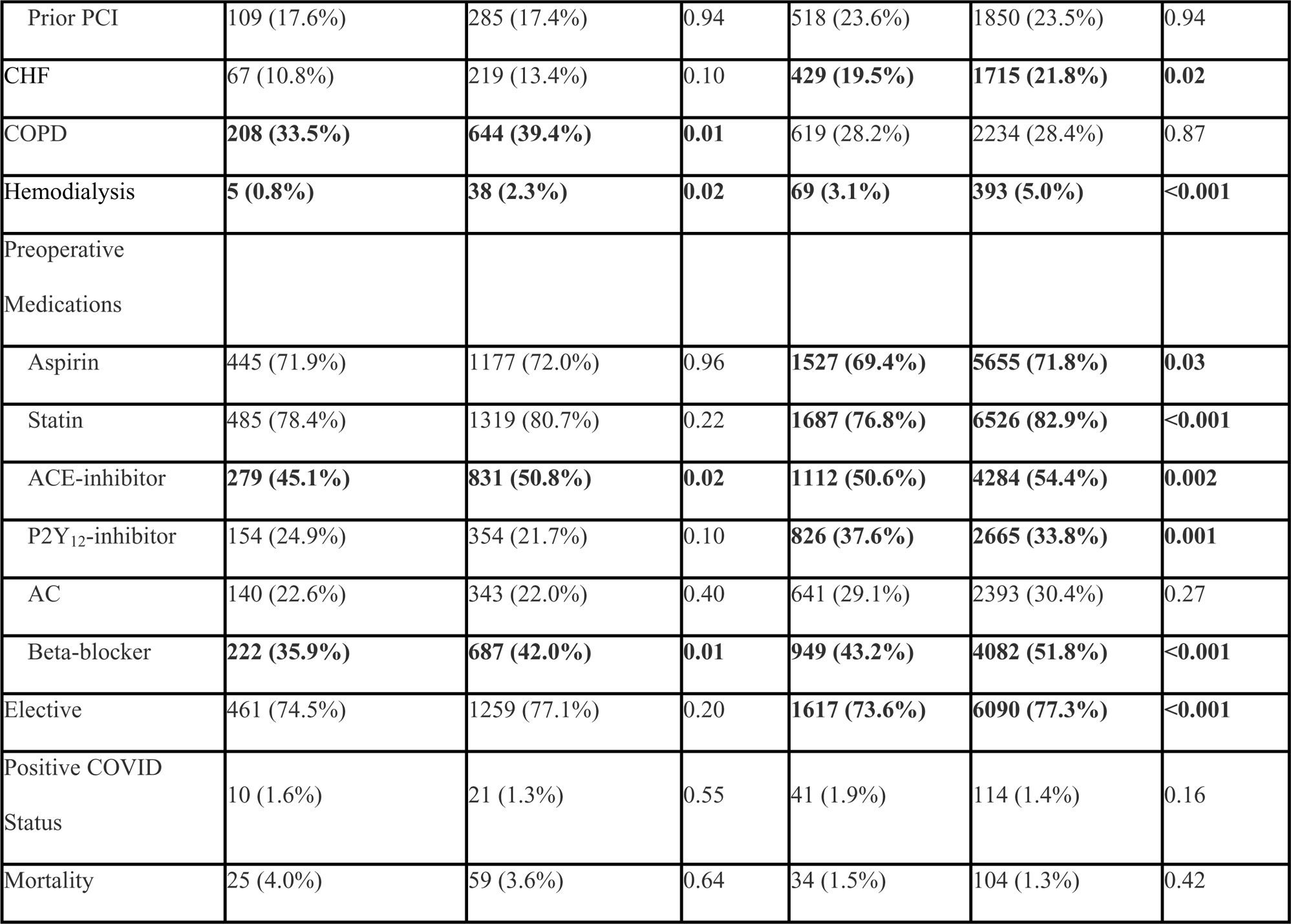
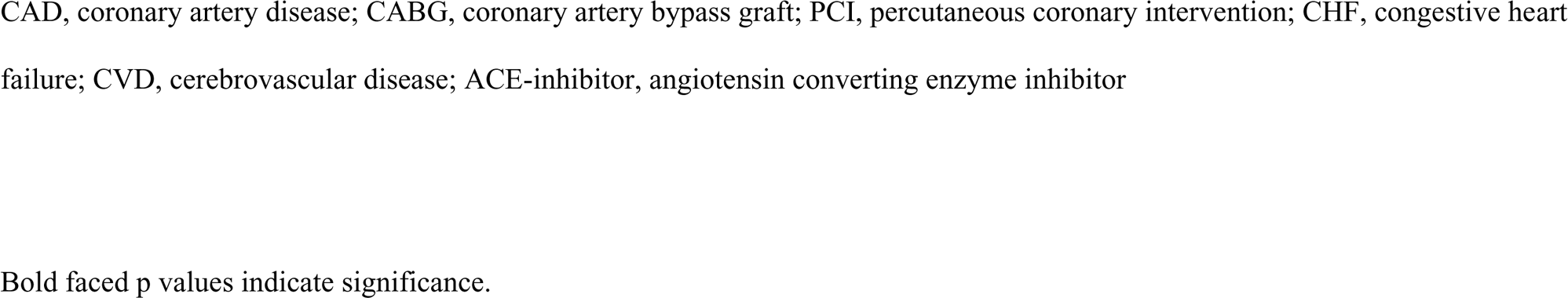
Baseline comorbidities and characteristics of unvaccinated and vaccinated patients undergoing revascularization for peripheral artery disease.

Unlike the overall study population or patients undergoing infra-inguinal bypass, white patients were over-represented in the unvaccinated cohort who underwent supra-inguinal bypass (82.9% vs 75.2%, p <0.001). Vaccinated patients who underwent peripheral revascularization were more likely to have a history of HTN, diabetes, and HD and to be on ACE-inhibitors or beta-blockers. In patients undergoing supra-inguinal bypass, there was no significant difference in the likelihood of undergoing an elective procedure between vaccinated and unvaccinated patients.

In patients with underwent either supra- or infrainguinal bypass, there was no significant difference in the proportion of vaccinated and unvaccinated patients with a positive COVID diagnosis and there was no significant difference in perioperative mortality.

## Discussion

In this retrospective analysis of a large, multicenter database, vaccination against COVID was significantly associated with reduced perioperative mortality after major vascular surgery. Despite presenting with a high burden of comorbid conditions, including hypertension, diabetes and coronary artery disease, vaccinated patients had reduced odds of perioperative mortality after open AAA, EVAR, CAS, and infra-inguinal lower extremity open revascularization procedures. Our results underscore the efficacy of the COVID vaccination in reducing perioperative all-cause mortality, particularly in older, high-risk patients.

Recently published reports have examined the protective effect of COVID vaccination amongst a different subset of surgical patients. In a retrospective, multicenter cohort review of patients undergoing high risk surgery, Sharath et al found that the unadjusted and adjusted 90-day mortality rates between vaccinated patients with and without a recent COVID-19 diagnosis were comparable. Moreover, in a subgroup analysis of matched vaccinated and unvaccinated COVID positive patients, vaccination reduced 90-day mortality by close to 50%.^15^ In a second, retrospective study of cardiac surgery patients, a cohort with similar risk factors to those undergoing vascular surgery, Blumenfeld et al examined the effect of perioperative COVID-19 infection on 30-day mortality before and after the development of the COVID vaccine. The authors reported a significantly elevated risk of perioperative mortality in COVID-19 positive patients prior to vaccine availability. However, after universal vaccination protocols were put in place, the association between COVID-19 infection and increased perioperative mortality was no longer apparent.^16^ Although further research is needed, it has been proposed that the reduced mortality may be attributed to effectiveness of the vaccine in quelling the systemic inflammatory state induced by virus which is further heightened in the context of surgery.^15^

The reduced all-cause mortality seen in our study population of largely COVID negative patients cannot be fully explained by the effectiveness of the vaccine in preventing COVID-19-related deaths. A similar reduction in all-cause mortality was observed in a population-based study out of Australia, in which the authors proposed several possible explanations for this finding, including indirect reduction in outcomes associated with COVID-19 infection, such as cardiovascular and pulmonary complications.^17^ Given that vascular surgery patients are amongst the highest risk patients for postoperative cardiac complications,^18^ this hypothesis seems reasonable. An additional consideration brought forth by the authors was the effect of confounding, particularly in the form of behavioral differences such as the healthy vaccinee effect.^17^ However, it is worth nothing that vaccinated patients within our study tended to present with more serious comorbid conditions which we believe reduces the likelihood that vaccination status is a simply a marker for access to and trust in the health care system.

The higher proportion of older and sicker patients who were vaccinated in our study is consistent with epidemiologic data within the United States. In the 2021, United States Census Bureau’s Household Pulse Survey of around 61,000 people, unvaccinated people were generally younger with 75% of the unvaccinated population under the age of 50.^19^ This pattern of vaccination can be at least partially attributed to the way the COVID vaccine was rolled out in most states with priority given to older adults and those living in group living facilities. However, beyond logistics, it is conceivable to believe that patients who are aware of serious comorbid conditions may be more likely to vaccinate for specifically that reason, given the widespread knowledge that people with these risk factors generally had worse outcomes with COVID infections. In fact, studies have described the relationship between increasing age or pre-existing medical conditions and the intention to vaccinate.^20–22^ The US Census Survey additionally reported a higher representation of non-Hispanic Black people within the unvaccinated cohort compared to the vaccinated cohort (13% vs 11%),^19^ a finding mirrored within our own study population. The significance of this disparity cannot be understated, particularly given the disproportionate impact of both COVID 19 and atherosclerotic cardiovascular disease on black individuals.^23^ A more comprehensive understanding of what drives vaccine hesitancy amongst vascular surgery patients should be undertaken to better address this issue.

During the height of the pandemic, major society guidelines suggested deferring elective procedures for several weeks after COVID-19 infection.^10^ However, in the endemic phase of COVID-19, as more patients have been exposed to the virus through infection and immunization, guidelines around surgical timing are in flux. The results of our study, in conjunction with others,^15,24^ suggests a protective effect of vaccination prior to surgery, particularly in patients whose risk factors predispose them to needing surgical intervention in urgent or emergent settings, such as those with vascular disease. However, further studies are needed to discern optimal timing between vaccination and operative intervention to provide greatest protection.

There are several limitations to this study, including its retrospective observational nature. While the use of the VQI-SVS database allowed us to capture a broader population, however, the registry lacks granular detail. For instance, the extent of vascular disease with regards to anatomy and morphology cannot be fully categorized. Furthermore, while data input is standardized, the accuracy and completeness of the information is center specific. Certain variables, including the length of time between vaccination and surgery, choice of vaccination, and implementation of booster shots were not consistently input into the database, limiting the extent of the analysis we were able to perform. Furthermore, the VQI does not generate data on the specific cause of death, so analysis was limited to all-cause mortality. Successful recovery from COVID infection confers a degree of immunity and it is unclear whether prior infection provides a synergetic, reductive, or independent effect on vaccination protection against perioperative mortality. Further longitudinal studies will be needed to elucidate these relationships.

## CONCLUSION

In our analysis of the Vascular Quality Initiative (VQI) database of all patients undergoing major vascular interventions, we found an association between vaccination status and improved perioperative survival. This association was most pronounced for patients undergoing AAA repair, carotid stenting and infrainguinal bypass. As the pandemic enters the endemic stage, future studies must focus on not only the prevention of viral infection in high-risk surgical patients, but also on downstream benefits that vaccines may impart on society.

## Data Availability

Data and analytic methods can be made available upon request pending approval by the Vascular Quality Initiative Research Advisory Committee.

## Acknowledgements

None

